# The Mean Unfulfilled Lifespan (MUL): A new indicator of the impact of mortality shocks on the individual lifespan, with application to global 2020 quarterly mortality from COVID-19

**DOI:** 10.1101/2020.08.09.20171264

**Authors:** Patrick Heuveline

**Author notes:** Corresponding author: Patrick Heuveline < >.

## Abstract

Declines in period life expectancy at birth (PLEB) provide intuitive indicators of the impact of a cause of death on the individual lifespan. Derived under the assumption that future mortality conditions will remain indefinitely those observed during a reference period, however, the intuitive interpretation of a PLEB becomes problematic when that period conditions reflect a temporary mortality “shock”, resulting from a natural disaster or the diffusion of a new epidemic in the population for instance.

Rather than to make assumptions about future mortality, I propose measuring the difference between a period average age at death and the average expected age at death of the same individuals (death cohort): the Mean Unfulfilled Lifespan (MUL). For fine-grained tracking of the mortality impact of an epidemic, I also provide an empirical shortcut to MUL estimation for small areas or short periods.

For illustration, quarterly MUL values in 2020 are derived from estimates of COVID-19 deaths in 159 national populations and 122 sub-national populations in Italy, Mexico, Spain and the US. The highest quarterly values in national populations are obtained for Ecuador (5.12 years, second quarter) and Peru (4.56 years, third quarter) and, in sub-national populations, for New York (5.52 years), New Jersey (5.56 years, second quarter) and Baja California (5.19 years, fourth quarter). Using a seven-day rolling window, the empirical shortcut suggests the MUL peaked at 9.12 years in Madrid, 9.20 years in New York, and 9.15 years in Baja California, and in Guayas (Ecuador) it even reached 12.6 years for the entire month of April.

Based on reported COVID-19 deaths that might substantially underestimate overall mortality change in affected populations, these results nonetheless illustrate how the MUL tracks the mortality impact of the pandemic, or any mortality shock, retaining the intuitive metric of differences in PLEB, without their problematic underlying assumptions.

## Introduction

For months, the numbers of deaths from the novel coronavirus disease 2019 (COVID-19) have become part of the daily news cycle the world over. Even when related to the population size in deaths per capita ratio, however, these numbers do not really provide any intuition for the magnitude nor the dynamics of the pandemic. Quite useful for between-population comparisons, the age standardization of these ratios does not make them more easily interpretable.

The period life expectancy at birth (PLEB) is probably the most readily interpretable of the period indicators mortality. Translating a number of deaths from a given cause into its impact on PLEB involves multiple steps but is fairly straightforward.^1, 2^ Unfortunately, the intuitive appeal of the PLEB, its interpretation as a measure of the individual lifespan, derives from the assumption that period mortality conditions will continue to prevail indefinitely. Declines in PLEB induced by a relatively rapid and likely temporary increase in mortality, such as currently experienced with the COVID-19 pandemic, can thus provide misleading indicators of changes in the individual lifespan.^3^

The assumption is even more problematic when PLEB or changes therein are estimated for smaller populations and shorter periods of time. Tracking the pandemic at a finer-grained geographical and temporal scale undoubtedly provides better insights on the pandemic than annual, national averages.^4, 5, 6^ But while the assumption underlying an annual PLEB estimate—that mortality conditions of a given year will be repeated year after year in the future—may seem unlikely, the seasonality of mortality makes the indefinite repetition of the mortality conditions in any fraction of a year not just unlikely but impossible.^7^ When referring to mortality conditions not only in a short period but also in a small area, the assumption on which PLEB estimates are build resort to a Groundhog-Day-like^8^ time loop repeating itself in a small area from which individuals are unable to leave.

For a both interpretable and scalable, over space and time, measure of the impact of changing mortality conditions on the individual lifespan, I suggest an alternative to the reduction in PLEB, the Mean Unfulfilled Lifespan (MUL). Making no assumption about future mortality, the MUL translates past changes in mortality into average difference in the length of lived lives. That difference is obtained by comparing the actual average age at death during a given period and the expected ages at death of the same individuals in the absence of mortality changes, whether induced by a specific cause of death or by an event affecting multiple causes of death. The MUL can thus be estimated for populations of any size and for periods of any length, from data on excess deaths or cause-specific deaths, as illustrated here with COVID-19 mortality data from 159 national populations and 122 sub-national populations in Italy, Mexico, Spain and the USA, for each sex and each quarter in 2020. The MUL also equals the product of (1) the proportion of deaths in the population during a reference period that are due to a specific cause or event and (2) the average reduction in length of life among individuals who died from that specific cause or due to that specific event in the population during the reference period. In the case of COVID-19, I show that for a given population the value of that average reduction only changes very slowly over time, providing an easy short-cut for fine-grained tracking of the pandemic.

### Conceptual Detour

Assessing PLEB reductions induced by a specific cause or event requires two period life tables, one representing the prevailing mortality conditions and another one representing the counterfactual mortality conditions expected in the absence of that cause or event. The assessment involves a relatively copious amount of life table manipulations, but decades ago Nathan Keyfitz provided most useful insights as to what these manipulations boil down to. Considering the related issue of estimating the increase in PLEB brought by the permanent elimination of a cause of death, he summarized that the increase “depends on the average time that elapses before the persons rescued will die of some other cause.”^9^ Conversely, the decrease induced by a new cause of death depends on the average time that would have elapsed before the persons who died from the new cause would have died from other causes.

This average time can be derived from the synthetic cohort approach modelled in the period life table where each death at age *a* from a cause *C, d*^*C*^(*a*), reduces the number of person-years lived by the life expectancy at age *a* in the absence of that new cause, *e*^*o-C*^(*a*). This assumes that persons dying from the new cause would have had the same life expectancy in the absence of that cause as same-age persons who survived that cause. This common assumption may appear unlikely, and interactions between causes of death can be incorporated instead, but the data requirements are substantial. Under the common assumption, the difference in PLEB is thus the average over all members of the synthetic cohort, *l*_0_ (the radix of the life table), of the difference in person-years lived by cohort members:

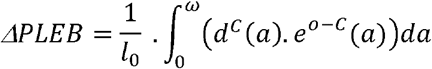

Keyfitz’ insight relates to the concept of “potential years of life lost”^10^ developed a couple of decades earlier still. The initial approach, designed to measure premature mortality, compared ages at death to a fixed value (70 or 75 years).^11^ This approach is not suited to study cause of deaths at older ages since deaths at ages above the fixed value are not considered.^12^ In burden-of-disease assessments, it has become customary to estimate Years of Life Lost (YLL) as:

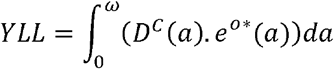

where *D*^*C*^(*a*) is the number of deaths from a certain cause *C* at age *a* observed in the population during a reference period and 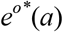 is life expectancy at age *a* in a counterfactual life table. YLL to COVID-19 have been estimated using this approach.^13, 14, 15^

Three differences between the two above equations can be observed. First, YLL are estimated from actual numbers of deaths by age rather than from numbers of life table decrements. This implies that the YLL estimate is sensitive to the age distribution of the population, as in turn it affects the distribution of deaths by age. While the PLEB is not an age-standardized measure,^16^ it equals the inverse of the “stationary” death rate, that is, a weighted average of the period age-specific death rates with weights derived from these death rates through life table construction. Using these internally-derived weights rather than an external, standard age distribution, the stationary death rate and PLEB are independent of the actual age composition of the population. This relative advantage of the difference in PLEB comes at the cost of using a “stationary” age distribution of deaths, however, represented by the life table decrements that result from indefinitely subjecting the population to the mortality conditions of the period. As discussed in the introduction, this assumption is precisely what is problematic for studying the impact of a mortality shock or an emerging disease such as COVID-19. Moreover, the actual distribution of deaths can be used in a population of any size and periods of any length, allowing for the mortality impact to be tracked on short temporal and small spatial scales for which interpreting differences in PLEB hardly makes sense.

The second difference refers to the counterfactual life expectancies. In global burden-of-disease assessments, a universal life table representing optimal survival conditions is typically used. This has the advantage of making YLL for different populations additive allowing for the derivation of a global estimate of YLL due to a cause by simple summation. However, using a universal life table may misrepresent the actual gains from averting a death in a specific population.

The last difference concerns the denominator, or lack thereof in YLL. The use of a denominator in the difference in PLEB allows for relating a total number of years in a population, measured by YLL, to a number of persons, and thus for a more intuitive interpretation as an average difference in years lived per person. Two ratios involving YLL can be found in the literature. First, the average YLL (AYLL) relates YLL to all the deaths from that cause in the population during the reference period, *D*^*C*^:

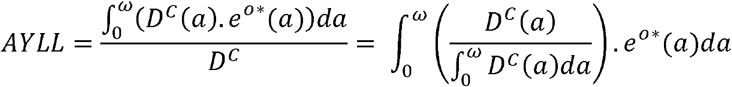

The AYLL thus represents the average (universal) life expectancy left to population members who died from the specific cause during a given period. On the one hand, it is a coherent measure as its denominator includes all the deaths that contribute to lost years in the numerator. On the other hand, it is only a function of the distribution of deaths by age, irrespective of the prevalence of that cause of death. The AYLL thus cannot provide a measure of the intensity of a mortality shock. The second measure is YLL per capita. While it does depend on the prevalence of the different causes of death, it is a less coherent measure as it includes in the denominator all the individuals in the population, including many that survived the mortality shock and do not contribute to YLL in the numerator. In turn, this complicates providing a precise interpretation for the value of YLL per capita.

Considering the advantages and limitations of the extant measures, I propose to add one measure of the mortality impact of a cause of death or an event on the individual lifespan. This measure, the Mean Unfulfilled Lifespan (MUL), is intended for situations where the underlying assumptions of PLEB might be implausible, and thus based, as the estimate of YLL, on actual numbers of deaths in a population during a period rather than on life table decrements. To retain its intuitive interpretation, however, the MUL is structured like the difference in PLEB as summarized by Keyfitz and, using counterfactual life expectancies representing the mortality conditions in the population of interest, similarly expressed as an average difference in person-years lived per person. Since the life table radix, *l*_0_, equals the sum of all decrements at all ages, the structural equivalence is maintained by defining the MUL as:

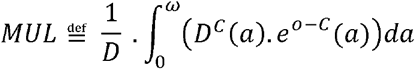

where *D* is the total number of deaths (from all causes at all ages) during the reference period. This intuitive interpretation of the MUL can be derived by rewriting this defining equation as:

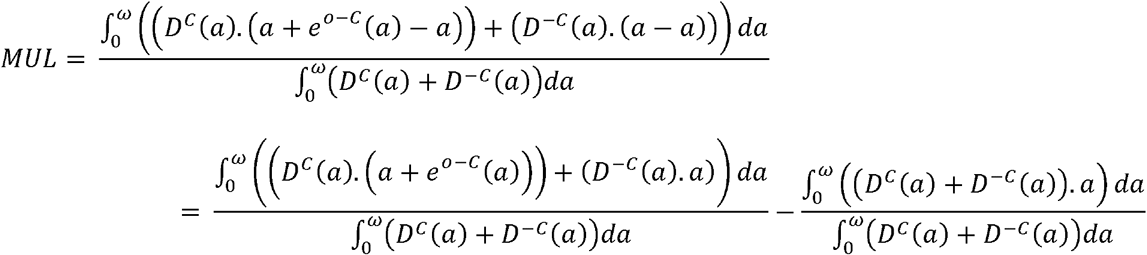

where *D*^*-C*^(*a*) is the number of deaths from all causes but *C* at age *a* observed in the population during the reference period. The second term represents the average age at death in a given period. If we assume that individuals who die of other causes than *C* die at the same age as they would have in the absence of cause *C* (no indirect effect of cause *C* on other causes of death), and that individuals who die of cause *C* at age *a* would have otherwise lived to age *a*+*e*(*a*), the first term represents the average expected age at death of the same individuals in the absence of cause *C*. The MUL is thus the difference between the average age at death in the population during a given period and the average expected age at death of the same individuals in the absence of cause *C*.

The assumption that cause *C* has no indirect effect on other causes of death is actually not required. As will be shown below, one can also derive the MUL from data on all deaths, and distinguishing between deaths that, based on counterfactual “benchmark” mortality conditions, were expected to occur in that period and those that were not, use the latter number of “excess” deaths instead of deaths from a specific cause. Finally, note that the MUL differs from changes in average ages at death across periods, which can be readily measured but may actually be positive with the emergence of a new cause of death if that cause affects people who are older on average that those dying from other causes. To sum up this conceptual detour, the MUL thus complements existing indicators of the impact on the individual lifespan of a cause of death by providing a measure of the average potential years of life lost to a specific cause of death, or to any mortality shock, for all population members dying in a certain period, regardless of their cause of death.

### Empirical Shortcut

Calculating the MUL for small geographical areas and short periods is not conceptually problematic since it captures the actual length of lives that ended there and then, unlike differences in life expectancies that require assumptions about future conditions. The demand on data (including a separate counterfactual life table for each population of interest) is substantial, however, and the life table manipulations are not particularly straightforward.

To simplify the estimation, the MUL can be rewritten as:

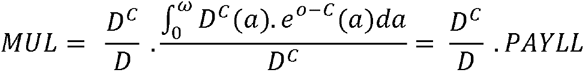

The second term is similar to the AYLL in burden-of-disease assessments, but again based on population-specific, counterfactual life expectancies instead of universal ones, and to underscore the difference is termed here the Population AYLL (PAYLL). In a given population, the PAYLL is a weighted average of counterfactual life expectancies that are estimated from prior conditions and do not change over time. Meanwhile, the weights are the ratios:

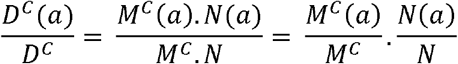

where *M*^*C*^ and *M*^*C*^(*a*) are the all-age death rate and the death rate at age *a* from a specific cause, and *N* and *N*(*a*) are the total population size and number of individuals at age *a* in the population. These weights should also be expected to vary little within short periods because their values depend on the population composition, *N*(*a*)/*N*, and on the age pattern of cause-specific death rates, *M*^*C*^(*a*)/*M*^*C*^, both of which should vary little within short periods. To confirm this, Figure 1 compares the distribution of provisional COVID-19 death counts in the USA at four points in time: on May 13 (one of the earliest dates for which this distribution is available) and on July 1^st^, September 30^th^ and December 30^th^.

**Fig 1:**
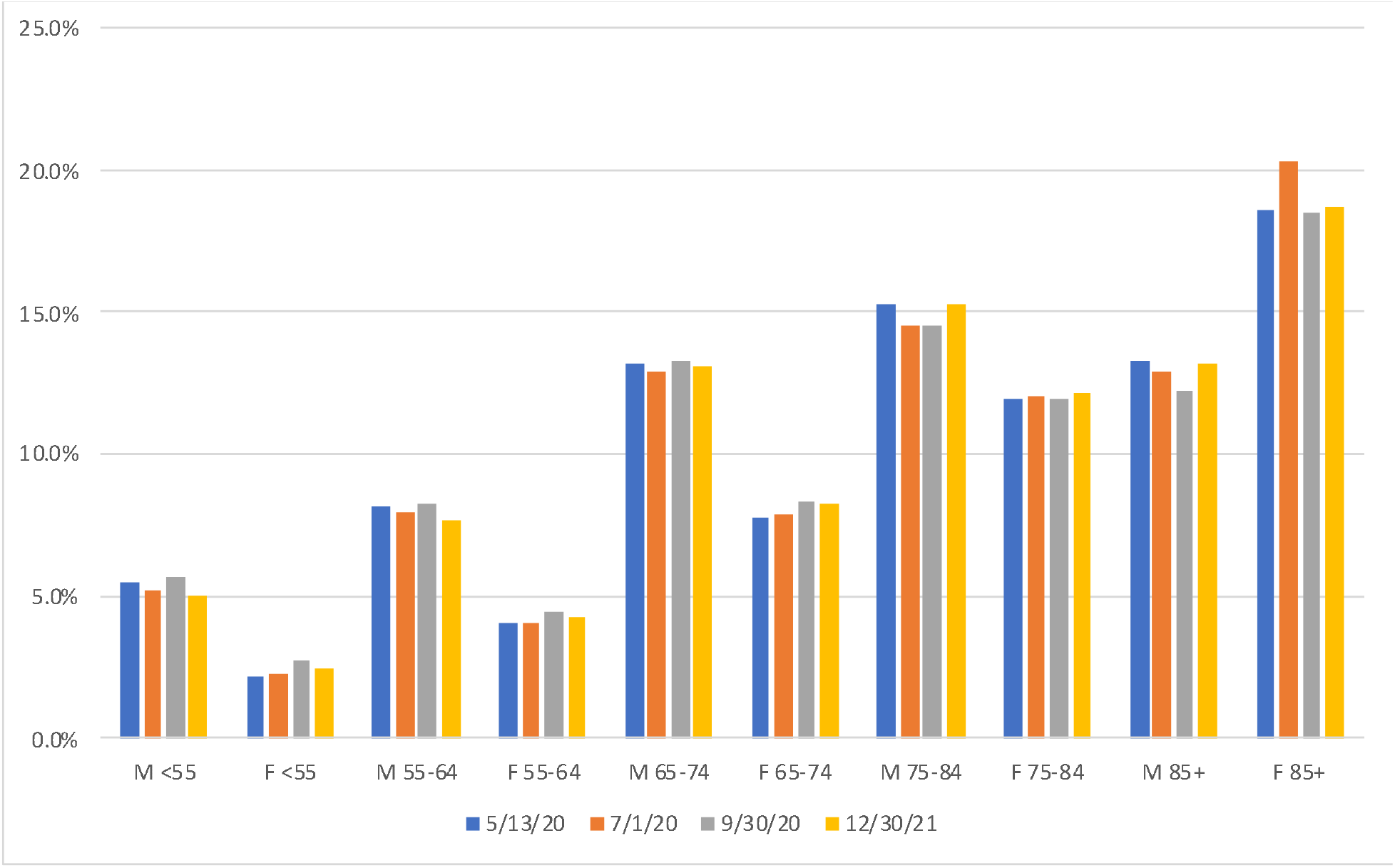
Distribution of Provisional COVID-19 Death Counts, by Sex- and Age-Groups, USA as of 5/13 (54,860 deaths), 7/1 (112,223 deaths), 9/30 (194,087 deaths) and 12/30 (301,671 deaths) Source: Centers for Disease Control and Prevention (CDC)

This suggests that the value of PAYLL can be expected to only change slowly over time and to be relatively close across populations with similar life expectancies and population compositions. In particular, MUL values for a sub-population or during a sub-period can be approximated as the product of the PAYLL for the whole population or the entire period and the variable all-age ratio of deaths from a specific cause to all deaths, *D*^*C*^/*D*, in the sub-population or during that sub-period. As noted above, this can be readily extended to any mortality shock, with data on excess deaths and all-cause deaths.

## Materials and Methods

The equations defining YLL and the MUL look deceivingly simple. To implement them, one has to apply estimates of life expectancies, which refer to exact ages, to numbers of deaths that are available or can be estimated only for age intervals. The varying value of life expectancy on a closed age interval is typically approximated by linear interpolation,^17^ in which case the contribution of the closed age interval between ages *x* and *x*+*n* to the MUL equals:

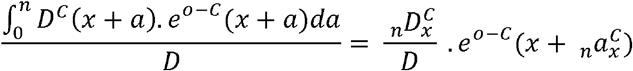

where _*n*_*D*_*x*_^*C*^ is the number of individuals between ages *x* and *x*+*n* who died of cause *C* during the reference period and _*n*_*a* _*x*_^*C*^ is the average number of years lived after age *x* by these individuals. In turn, life expectancy at exact age *x*+ _*n*_*a* _*x*_ ^*C*^ can be derived by linear interpolation between the values of lifeexpectancy at ages *x* and *x*+*n*.

The linear interpolation is more problematic for wider the age intervals and for older age groups. In the 2018 US life table for males for instance,^18^ life expectancy declines by 2.9 years between ages 75 and 80 and by 2.4 years between ages 80 and 85. In this case, the linear approximation over-estimate life expectancy in the interval. The average number of years lived after age 75 by individuals dying between ages 75 and 85 is 5.4 years, and linear interpolation would yield a life expectancy at age 80.4 years of 8.5 years, whereas life expectancy has already dropped to 8.4 years at age 80. This upward bias is particularly undesirable in the case of COVID-19 deaths at those ages because COVID-19 victims are more likely to suffer from other long-term conditions that more likely reduced their life expectancies compared to same-age individuals. Even more problematic with linear interpolation is the open-ended interval, as it requires that an arbitrary upper age limit be set. Unfortunately, these couple of issues may concern a large share of COVID-19 deaths: as shown in Figure 1, close to 60% of US deaths are above age 75 years and reported in just one ten-year closed interval (75 to 84 years) and one open age interval (over 85 years).

In this respect, working with all deaths in a given period rather than with deaths from a specific cause of interest presents an important empirical advantage in addition to the benefit of assessing both the direct and the indirect effects of that cause of death. Using all deaths in a closed age interval between ages *x* and *x*+*n*, _*n*_*D*_*x*_, one may first calculate the contribution to the MUL of the deaths that were expected to occur under the counterfactual mortality conditions, _*n*_*D*_*x*_ ^-*C*^. For these deaths, the difference in length of life averages to the difference between the average number of years lived in the age interval under the prevailing conditions, _*n*_*a*_*x*_, and under the counterfactual conditions, _*n*_*a*_*x*_ ^*-C*^. The advantage of this approach is that setting an arbitrary upper value for the open-ended age interval becomes unnecessary because, having already reached age *N*, individuals who contributed to the number of deaths in the open age interval would all have been expected to die in the same open-ended age interval under the counterfactual mortality conditions, albeit possibly not in that period. For all the open-interval deaths over age *N, D*_*N+*_, the difference in length of life thus averages to the difference between life expectancy at age *N* under the prevailing conditions, *e*^*o*^(*N*), and under the counterfactual conditions, *e*^*o*-*C*^(*N*).

This approach limits the issue of estimating the average reduction in length of lived lives on closed age intervals from values of life expectancies that vary on these age intervals to “excess” deaths, _*n*_*D*_*x*_ - _*n*_*D*_*x*_^-*C*^. An alternative to linear interpolation derives from the fact that an individual’s expected length of life (*e*^*o*^(*x*)+*x*) gradually increases with age *x*. Life expectancy at any age is thus larger than the difference between life expectancy at an earlier age and the difference between the two ages:

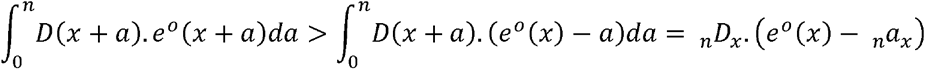

Applying this approximation to excess deaths on any closed age interval will induce some underestimation of the average life expectancy of individuals dying on the interval, which appears preferable in this case to the overestimation induced by linear interpolation.

Adding the contributions of the different types of deaths yields:

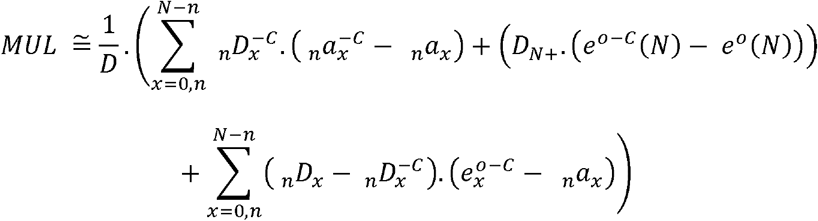

Rearranging the sums corresponding to the closed age intervals and using the multiple-decrement life table relationship *e*^*o*^(*N*)/ *e*^*o*-*C*^(*N*) = *D*^*-C*^_*N+*_/*D*_*N+*_, this can be rewritten as:

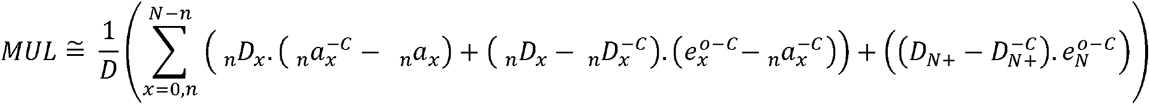

Using this approximation to estimate the impact of COVID-19 on the individual lifespan in each quarter of 2020 first requires a life table representing survival conditions in 2020 in the absence of COVID-19 whose values of *e*_*x*_ ^*o-C*^ and _*n*_*a*_*x*_ ^-*C*^ can be used. Combined with the number of individuals by sex and age-group, the life table values of age-specific death rates, _*n*_*m* ^-*C*^, then provide the expected numbers of deaths _*n*_*D*_*x*_ ^-*C*^ in the absence of COVID-19. Population data and life table functions for countries were obtained from the UN Population Division.^19^ Corresponding data for sub-populations in Italy, Spain, and the US were obtained from national statistical agencies.^20, 21, 22, 23^

New life tables representing actual mortality conditions (with COVID-19) in each quarter must then be derived to calculate the corresponding values of _*n*_*a*_*x*_. The construction of these life tables requires the quarterly numbers of deaths by sex and age-group. In countries where vital statistics are incomplete not available yet, but estimates of COVID-19 deaths are available, the total number of deaths, _*n*_*D*_*x*_, can be obtained by adding these estimates (through a multi-decrement life table to adjust for competing risks of deaths, which typically assumies no indirect effects on other causes of death)^24^ to the expected numbers of deaths in the absence of Covid-19 (_*n*_*D*_*x*_^-*C*^). When COVID-19 estimates are not broken down by sex and age-group, an alternative is to use a reference set of age-and-sex death rates from COVID-19 from another population for which these rates are deemed reliable.^25^ Centers for Disease Control and Prevention (CDC) data provided the reference set of age-and-sex death rates from COVID-19.^26^ Estimates of COVID-19 deaths by March 31, June 30, September 30 and December 31, 2020 were taken from the IHME.^27^ All of these data were downloaded from institutional websites.

## Results

Figure 2 shows MUL values for selected national and sub-national populations in each of the four quarters of 2020 (for both sexes combined). First-quarter MUL values illustrate substantial impacts of COVID-19 on longevity already in parts of Spain (Madrid, 3.26 years) and Italy (Lombardy 2.76 years). In the second quarter, the impact continued to increase in these two nations, with the MUL reaching 4.32 years in Catalonia for instance, but also in other European nations such as Belgium (2.80 years). However, the second-quarter MUL values reached their highest values in parts of the USA (New Jersey, 5.56 years, and New York, 5.52 years). Very high second-quarter values are also estimated for parts of South America (Ecuador, 5.12 years) and Mexico (Baja California, 5.10 years).

**Fig 2:**
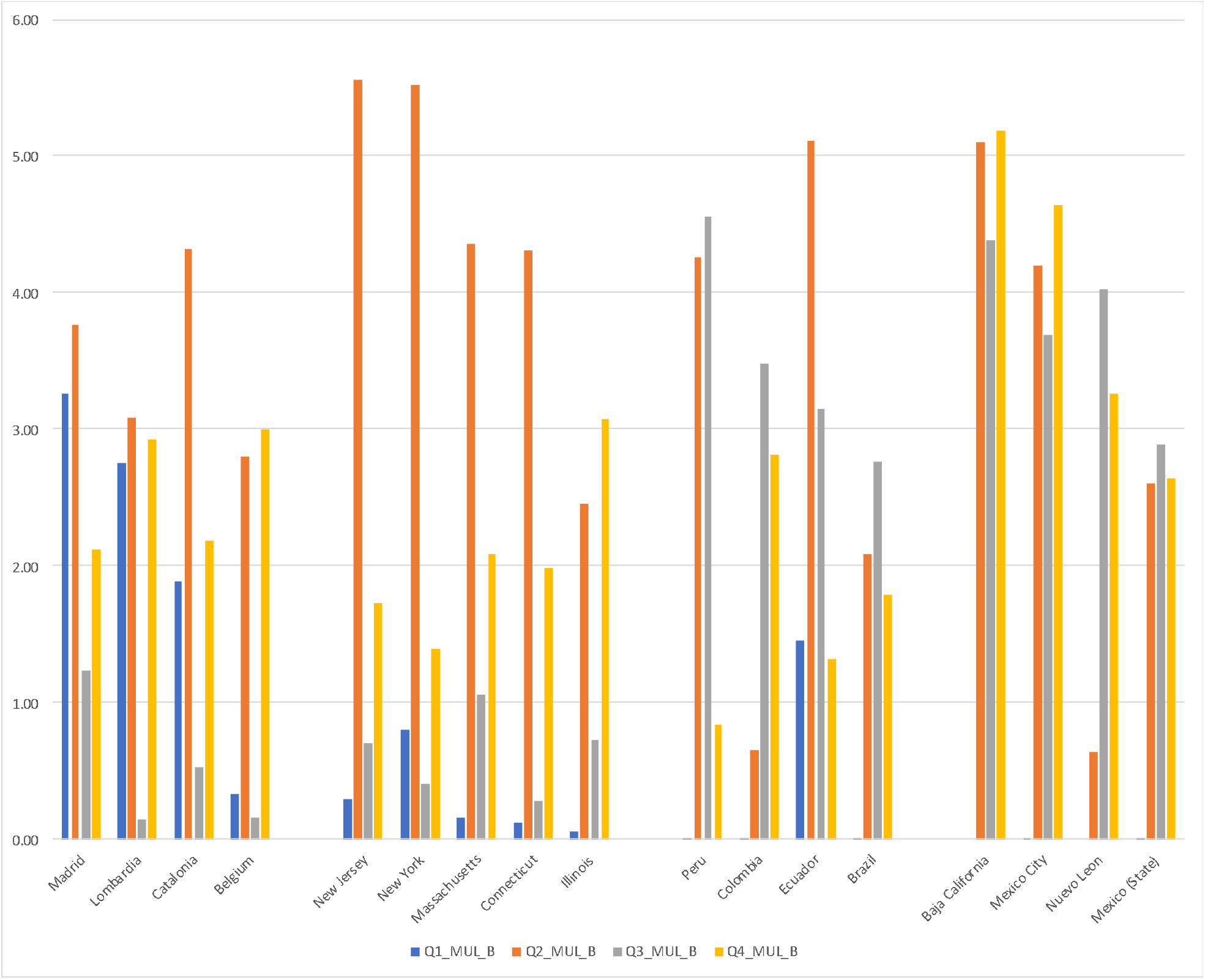
Quarterly Mean Unfulfilled Lifespan (MUL) for both sexes, in years, selected populations (populations with 5,000 or more COVID-19 deaths by January 1^st^, 2021, and the highest annual MUL values for 2020).

In the third quarter, MUL values continue to increase or remain high in parts of South America (Peru, 4.56 years) and Mexico (Baja California, 4.39 years, and Nuevo Leon, 4.03 years), whereas they were declining in Europe and the USA. In the last quarter of 2020, MUL values decline in South America, whereas the highest values remain for parts of Mexico (Baja California, 5.19 years, and Mexico City, 4.64 years). Fourth-quarter MUL values also rebound in parts of Europe (Belgium, 3.00 years) and the USA (Illinois, 3.07 years). MUL values for men are typically higher than for women, and the average age at death of men who died in New Jersey during the second quarter was nearly 6 years younger than their average expected age at death without COVID-19 (5.97 years, compared to 5.14 years for women, full results shown in supplementary file, Table S1).

The estimated quarterly PAYLL values (see supplementary file, Table S1) can be used to apply the suggested empirical short-cut. These values range from a low of 9.7 years in Bulgaria to a high of 26.4 years in Qatar. These differences can be explained by different age compositions, with younger compositions giving more weight to remaining life expectancies at younger ages, which are obviously higher. As expected, however, these values change relatively little from one quarter to the next for a given population. Figure 3 shows peak MUL values for a rolling seven-day period, derived from quarterly PAYLL values, were reached between mid-March and the end of June in Madrid (9.12 years on 3/29), in New York (9.20 years on 4/13) and in Baja California (9.15 years on 6/9).

**Fig 3:**
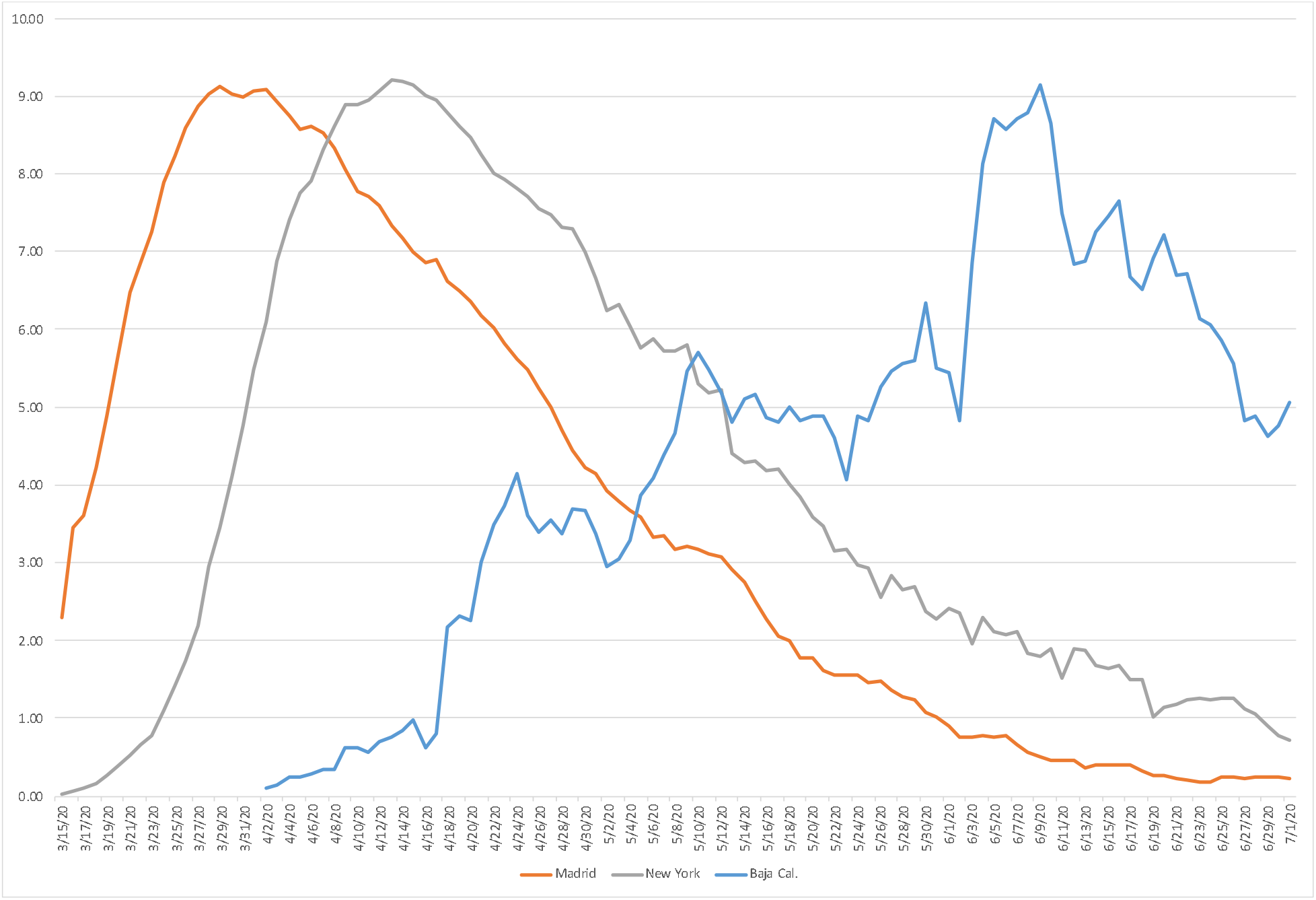
Mean Unfulfilled Lifespan (MUL) for both sexes, by rolling seven-day periods, in years.

The same approximation but for sub-populations is illustrated by focusing on the province of Guayas in Ecuador. Data on the monthly number of deaths by province show the marked increase in March, April and May from a baseline of 1,700-2,000 per month in January, February and again in June.^28^ In April, the number of deaths reached 12,004, of which the January-February-June average suggests only 17.2% might be estimated to be from causes other than COVID-19 (without adjustment for competing causes). No specific life table is available to estimate MUL values for the province directly, but based on the second-quarter PAYLL derived for Ecuador, individuals appeared to have died 12.6 years younger, on average, that their expected age at death in April in the province of Guayas.

## Discussion

The MUL is proposed here as an alternative to induced changes in PLEB to assess the impact of a cause of death of the individual lifespan in situations where the assumptions underlying life table construction are implausible, invalidating the usual interpretation of the PLEB. Complementing existing measures such as the AYLL and the YLL per capita, the MUL is similarly based on estimating the number of potential years of life lost corresponding to deaths from a specific cause or excess deaths from a specific event in a given period. As shown above, by averaging estimated potential years of life lost over the total number of deaths in the period, the MUL is structured like a difference in PLEB. The MUL retains the intuitive interpretation of a difference in PLEB as a change in average length of life, but for an actual cohort of individuals (those dying in the period) subjected to prevailing mortality conditions rather than for a synthetic cohort subjected to indefinitely constant conditions.

To illustrate the derivation and interpretation of the MUL in the context of COVID-19 mortality, MUL values were derived for a total of 281 populations for each sex and each quarter of 2020. As argued above, the MUL is best estimated from data on numbers of excess deaths by age and sex in a given period, estimated from data on all-cause mortality. At this writing, however, these data are not available from the vast majority of the national and subnational populations around the world. Instead, total numbers of quarterly COVID-19 deaths in each of these populations were used and their age-and-sex breakdown derived with the assumption of a shared age-and-sex COVID-19 mortality “pattern” (i.e., the same ratio of age-and-sex-specific death rates to the all-age, both-sex death rate in all populations). Excess deaths by age and sex were then derived through life table adjustments for competing causes of death (as COVID-19 deaths reduced exposure to other causes of deaths).

The adjustment for competing causes of death, as well as the estimate of potential years of life lost corresponding to each excess death is based on the assumption that, in the absence of a certain cause of death, individuals who died from that cause would have faced the same risk of death from other causes as any individual of the same age and sex. As discussed above, this standard assumption of YLL-based measures is problematic in the case of COVID-19 due to the higher proportion of several underlying long-term conditions (e.g., obesity) observed among COVID-19 victims. Adjusting for differences in long-term conditions prevalence is very data demanding, however, and the impact might not be as large as expected, a study finding that performing this adjustment reduces the AYLL for COVID-19 in the United Kingdom was from 13 to 12 years (average for both sex).^29^ An alternative strategy was proposed here that underestimates the value of potential years of life lost over an age interval by estimating its value at the beginning of the age interval. As shown in the supplementary files, this yields quarterly PAYLL values for the United Kingdom that vary between 11.9 and 12.0 years (Table S1), values thus quite close to what the adjustment for underlying long-term conditions might have provided. While there is of course no guarantee that this strategy would apply equally well to causes of death other than COVID-19, ignoring comorbidities will likely lead to some overestimation of AYLL for most causes of death, and purposely underestimating the number of potential years of life lost will likely be preferable in other situations as well.

The MUL estimates derived here can also be improved by using actual age-and-sex breakdowns of COVID-19 deaths in populations of interest, which are becoming available for an increasing number of nations.^30^ Here too, however, the actual impact on MUL estimates might not be as large as expected. The age patterns of CoViD-19 death rates available so far similarly exhibit remarkable regularities, with some modest variation in the slope of these age patterns at the oldest ages, probably due to the number of fatalities in nursing homes across Europe and the USA.^31^ Considering the two countries with the largest number of COVID-19 deaths in 2020, substituting the actual sex and age pattern in Brazil to the one derived by assuming the same mortality pattern as in the USA only changes the estimated reduction in 2020 PLEB for Brazil from 1.72 to 1.67 years.^32^ The structural similarity between the MUL and differences in PLEB suggest than using population-specific data on COVID-19 mortality by age and sex might only improve MUL estimates by a similarly modest order of magnitude.

To date, by far the strongest reason to caution against taking these MUL estimates at face value originates in lacking or incomplete data on the total number of deaths in 2020 for many populations and how this number might eventually relate to the number of reported COVID-19 deaths. Even in countries with reliable vital statistics, annual deaths data might not be finalized until several months into the following year. Moreover, the estimation of excess deaths is performed by comparison with benchmark mortality rates to represent counterfactual mortality conditions, and the results can be quite sensitive to the choice of a benchmark period (e.g., the last five years) and the estimation methodology (e.g., assumptions about mortality trends).^33^ In a study of 21 high-income nations,^34^ the official number of COVID-19 deaths from mid-February to end of May was found to be lower than the lower bound of the 95% credible interval for excess deaths during that period in 5 countries. The discrepancy was highest in Spain, with the credible interval from 47% to 91% more excess deaths than reported COVID-19 deaths. In the USA, between January 26^th^ and October 3^rd^, excess deaths were also estimated to be 38% higher than the COVID-19 death tally over the same period.^35^ Unfortunately, these ratios exhibit insufficient empirical regularities across populations (and possibly over time) to uniformly adjust numbers of reported COVID-19 deaths across populations for which the number of excess deaths remains unknown at this point.

More generally, another advantage of the MUL is that it remains interpretable regardless of its temporal and geographical scale, pertaining to deaths in a given population in a given period, whereas we argued the PLEB becomes difficult to interpret as a short-term, micro-level indicator. With the PAYLL values provided in supplementary file (Table S1), MUL values can easily be approximated as the product of the corresponding PAYLL value and the ratio of COVID-19 to total deaths in the population during a given period. The approximation can be used for short-duration periods as long as PAYLL can be assumed to be almost constant over time and quarterly PAYLL estimates are found indeed to change little from one quarter to the next. This approximation can also be used for sub-populations for which the necessary data are only provided of the entire population, but only when the age compositions of the sub-populations can be held as relatively similar.

This relates more generally to the fact that, like YLL-based measures, the MUL is not sex- or age-standardized and MUL comparisons across populations that differ markedly in age composition will be biased. On the one hand, all else equal, a younger population composition yields a younger distribution of deaths and a higher PAYLL value. On the other hand, known variations of COVID-19 mortality with age^36^ suggest that an older population distribution contributes to a higher proportion of COVID-19 deaths relative to all deaths. Only by accident would the two, opposite age-composition effects cancel each other and in general the MUL is not a standardized measure. This represents one disadvantage compared to differences in PLEB. However, this advantage over the MUL again comes at a cost, since the internal derivation assumes the period mortality conditions will become permanent.

When this assumption is not tenable, the MUL provides an unstandardized alternative to differences in PLEB, similarly structured as an average difference in length of lives lived per person. Related to other unstandardized measures such as the AYLL or YLL per capita, the MUL’s interpretation pertains to an actual death cohort, that is, population members who died during a certain period rather than to a synthetic cohort as represented in the life table. To reiterate, the MUL indicates the difference between their average age at death and their average expected age at death had a temporary mortality shock not occurred, and its measurement requires no assumption that these temporary conditions will either pass entirely or extend indefinitely into the future.

## Data Availability

All input materials are from online data sources listed in references list.
Full results provided as supplementary information.

## Acknowledgments

The author benefited from facilities and resources provided by the California Center for Population Research at UCLA (CCPR), which receives core support (P2C-HD041022) from the Eunice Kennedy Shriver National Institute of Child Health and Human Development (NICHD). The author thanks Michel Guillot, Philippe Bocquier, Tim Riffe, Michel Garenne, Christophe Guilmoto and Sam Preston for comments on an earlier draft of this manuscript.

